# Wildfire smoke exposure and early childhood respiratory health: a study of prescription claims data

**DOI:** 10.1101/2022.09.20.22280129

**Authors:** Radhika Dhingra, Corinna Keeler, Brooke S. Staley, Hanna V. Jardel, Cavin Ward-Caviness, Meghan E. Rebuli, Yuzhi Xi, Kristen Rappazzo, Michelle Hernandez, Anne Chelminski, Ilona Jaspers, Ana G. Rappold

## Abstract

Wildfire smoke is associated with short-term respiratory outcomes including asthma exacerbation in children. As investigations into developmental wildfire smoke exposure on children’s longer-term respiratory health are sparse, we investigated associations between developmental WSE and first use of respiratory medications. Prescription claims from IBM MarketScan Commercial Claims and Encounters database were linked with wildfire smoke plume data from NASA satellites based on Metropolitan Statistical Area (MSA). A retrospective cohort of live infants (2010-2016) born into MSAs in six western states, having prescription insurance, and whose birthdate was estimable from claims data was constructed (N=184,703); of these, gestational age was estimated for 113,154 infants. MSA, gestational age, and birthdate were used to estimate average weekly smoke exposure days (*smoke-day*) for each developmental period: three trimesters, and two sequential 12-week periods post-birth. Medications treating respiratory tract inflammation were classified using active ingredient and mode of administration into three categories: : ‘upper respiratory’, ‘lower respiratory’, ‘systemic anti-inflammatory’. To evaluate associations between wildfire smoke exposure and medication usage, Cox models associating smoke-days with first observed prescription of each medication category were adjusted for infant sex, birth-season, and birthyear with a random intercept for MSA. Smoke exposure during postnatal periods was associated with earlier first use of upper respiratory medications (1-12 weeks: hazard ratio (HR)=1.094 per 1-day increase in average weekly smoke-day, 95%CI: (1.005,1.191); 13-24 weeks: HR=1.108, 95%CI: (1.016,1.209)); sex-specific HRs varied by post-birth exposure window. Protective associations were observed during gestational windows for both lower respiratory and systemic anti-inflammatory medications; it is possible that these associations may be a consequence of live-birth bias. These findings suggest wildfire smoke exposure during early postnatal developmental periods impact subsequent early life respiratory health.

## Introduction

Over the past two decades, frequency and severity of wildfire events has increased, resulting in greater land area being burned each decade (3.3 and 6.8 million average acres per year in 1990s and 2010s, respectively(1)) and substantial economic impact ($347 billion in 2016) that is expected to grow as wildfire frequency increases (2). Wildfire smoke is a complex chemical mixture of both gases and small particles and a major contributor to particulate matter (PM), including the fine fraction of PM (PM_2.5_)(3) which has been implicated in over 8.8 million premature deaths worldwide (4).

Particulate matter (PM) exposure during wildfires is associated with acute respiratory and other outcomes in children (5); in addition, wildfire exposures may increase exacerbation-of-asthma events as indicated by hospitalizations, emergency department visits, and outpatient visits (6). In adults, controlled exposure studies of short-term exposures to woodsmoke have underscored the role of smoke exposure in respiratory dysfunction, and provide evidence of both systemic and respiratory inflammation (7). In studies of biomass – a primary fuel source for wildfires – burning and woodsmoke, exposure to these pollutants was associated with asthma related symptoms including wheeze in children (8). Epidemiologic studies (9–14) provide further evidence that wildfire, woodsmoke, and biomass smoke exposures are associated with an increased risk of respiratory infection and reduced lung function in children (15,16) and adults (17–19).

While the acute effects of wildfire or biomass burning exposure have been somewhat established, there is little known about the effects of *in utero* or in early life wildfire particulate matter exposure on longer term respiratory health outcomes. Lung development continues through gestation into the postnatal period, a likely critical window of respiratory susceptibility to air pollution (20). Additionally, PM exposure during gestation (e.g., 21,22) and early childhood (e.g., 23) is linked to sex-specific respiratory health outcomes in humans (21). Additional evidence comes from recent studies of non-human primates where exposure to wildfire smoke in the post-natal period resulted in sex-specific attenuation of host-defense mediators and impaired lung function in adolescence (24).

As similar respiratory effects may also be possible in humans, we consider the research question: Does exposure to wildfire smoke during gestation or in the early postnatal period result in earlier first use of anti-inflammatory respiratory medications in early childhood? By investigating the association of smoke exposure during multiple developmental periods with first use of respiratory medication, we identify critical windows of development where exposure to wildfire smoke may result in increased respiratory vulnerability in the population of young children.

## Methods

### Population

We constructed a retrospective cohort of infants from the IBM MarketScan® Commercial Claims and Encounters Research Database (MarketScan) born into six western U.S. states. MarketScan is a proprietary deidentified claims database, comprised of data from private U.S.-based insurance companies. This private claims dataset includes hospitalizations, outpatient visits, services, and prescription claims data.

Children born between January 1, 2010 and December 31, 2016, in metropolitan statistical areas (MSAs) in California, Oregon, Washington, Idaho, Montana, or Nevada (**Figure 1**; **Table S1**) were eligible. Utilizing a validated algorithm (25,26) and ICD-9 or ICD-10 codes, birthdates were estimated for each live birth (**Appendix 1, Supplementary Material)**. The MSA was the smallest geographic unit available in MarketScan, and therefore the primary spatial unit for assessing wildfire smoke exposure.

**Figure 1.**
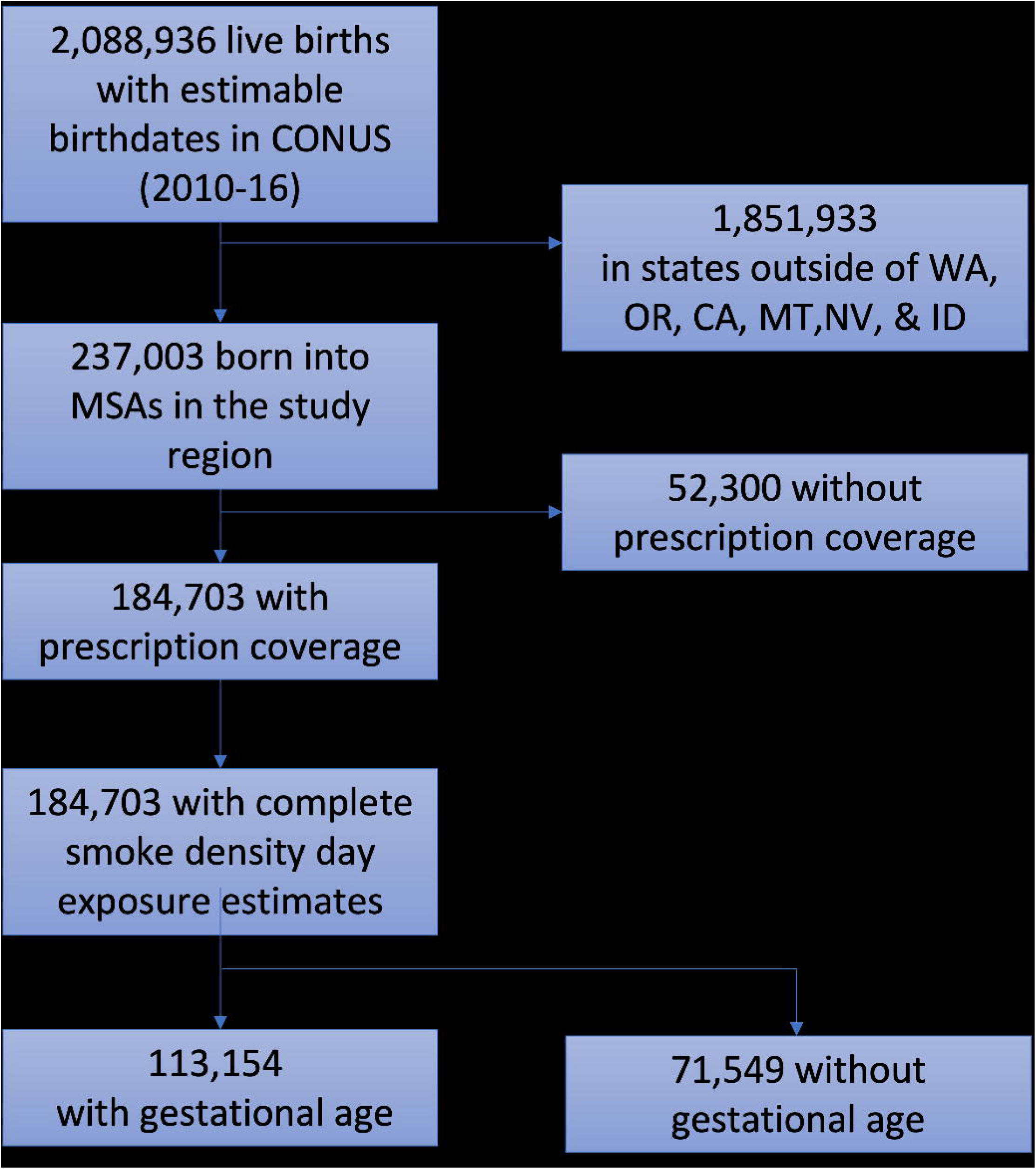
Cohort construction. WA, OR, CA, MT, NV, and ID refer to the states of Washington, Oregon, California, Montana, Nevada, and Idaho, respectively. MSA is Metropolitan Statistical Area and CONUS is Continental United States.

Birthdate and MSA were required to estimate exposure to wildfire smoke for all observations.

Each child was required to have continuous enrollment with the insurer with at least 1 week of prescription coverage starting at birth. For each child, follow-up begins after risk period; specifically, follow-up begins at birth for prenatal exposures, and after the post-natal risk period for postnatal exposures. For each outcome, the end of follow-up, measured in weeks, was either date of the outcome, the end date of continuous enrollment, or December 31, 2018, whichever came first. This resulted in two analytic cohorts: the full cohort with 182,387 liveborn children and a sub-cohort with 113,154 liveborn children that also had estimates for gestational age (hereafter, *GA sub-cohort*). This study (IRB No:20-2817) was evaluated and approved by the Institutional Review Board at the University of North Carolina at Chapel Hill.

### Gestational age estimation

Gestational age was used in the GA sub-cohort to estimate with greater precision the average weekly number of smoke exposure days for the portion of each trimester completed. For example, if a child had a GA of 34 weeks, T1 and T2 estimates would be calculated as described above, but T3 would be the average weekly smoke exposure from the 27^th^ week to the end of the 34^th^ week.

As the MarketScan dataset does not explicitly contain variables for gestational age (GA) at delivery, infant records were linked, where possible, to the birthing parent to allow for GA determination. To estimate GA at birth, we used an algorithm based on ICD-9 codes for the number of completed weeks of gestation, as described in **Appendix 2 (Supplementary Material)** (Adapted from 27); these codes were obtained either from the infants’ or the birthing parents’ records. Observations with GA were not evenly distributed across time due to the shift from ICD-9 to ICD-10 codes in the fall of 2015. Due to marked changes in GA coding between ICD-9 and ICD-10 codes, we chose to estimate GA based on ICD-9 codes only as they were used for the majority of the study period.

### Outcome: First use of prescription respiratory medication

Three outcomes were considered: the first prescription use of (1) upper respiratory, (2) lower respiratory, and (3) systemic anti-inflammatory medication. Classifications for each of the three outcomes are briefly summarized in **Table S2**. As systemic anti-inflammatory medications are often used in acute cases of inflammation or infection that could involve the lower respiratory tract, the upper respiratory tract or other organ system, we chose to analyze them separately from medications used more specifically for the upper or lower respiratory tract. If a child was prescribed medications from two outcome classes on separate or the same claim date, they were assumed to have the event in analysis of both outcomes. Medications were selected using a combination of *mode of administration* (*MSTFMDS* variable in MarketScan), *therapeutic drug classes* (*THERCLS*), and *therapeutic drug groups (THERGRP)* as defined in Micromedex RedBook (28) in consultation with a pediatric allergist, and referencing Up-To-Date (29); final classification of medications were validated by two physicians. The outcome event was defined as date of the first fill of prescription respiratory medication in drug claims data.

### Exposure: Average weekly wildfire exposure smoke-days

Wildfire smoke exposure data were obtained from the publicly available NASA satellite imagery-based Hazard Mapping System Fire and Smoke product (30,31) and were previously used to generate smoke-day exposure estimates for each ZIP Code Tabulation Area (ZCTA) in the study area (10). These data use visible plumes observed via satellite imaging as an approximation of true wildfire smoke exposure. Using these data, we constructed MSA-level daily smoke-day exposures as follows: First, we used the official crosswalk files released by the United States Office of Housing and Urban Development to define membership within an MSA for each ZCTA in the study area, for each year of the study (32,33). A MSA was defined as having a smoke-day on a given date if at least 25% of the population of the MSA experienced a smoke-day, based on weighting the binary ZCTA smoke-days by year-specific ZCTA population; we refer to this exposure measurement as the “25% threshold” smoke-day.

In sensitivity analyses, we further examined how estimated proportion of the MSA population exposed in developing the exposure metric might impact observed associations. Because each MSA contained a differing number of ZCTAs, we also defined two additional exposure measures for use in this sensitivity analyses: (1) if any ZCTA within that MSA had a smoke-day on a given date, the MSA was defined as having a smoke-day on that date [hereafter referred to as “0% threshold”]; and (3) if 50% of the population of the MSA experienced a smoke-day on a given date, based on weighting the binary ZCTA smoke-days by year-specific ZCTA population, the MSA was defined as having a smoke-day on that date [50% threshold]. Thus, the “0% threshold” and “50% threshold” roughly correspond to average weekly smoke-days experienced by greater than 0% or 50% of the MSA’s population, respectively (**Figure 2**). All analyses were conducted using each of these exposure thresholds, with the 25% threshold smoke-day exposure utilized in the primary analyses, and the 0% and 50% thresholds applied in sensitivity analyses.

**Figure 2.**
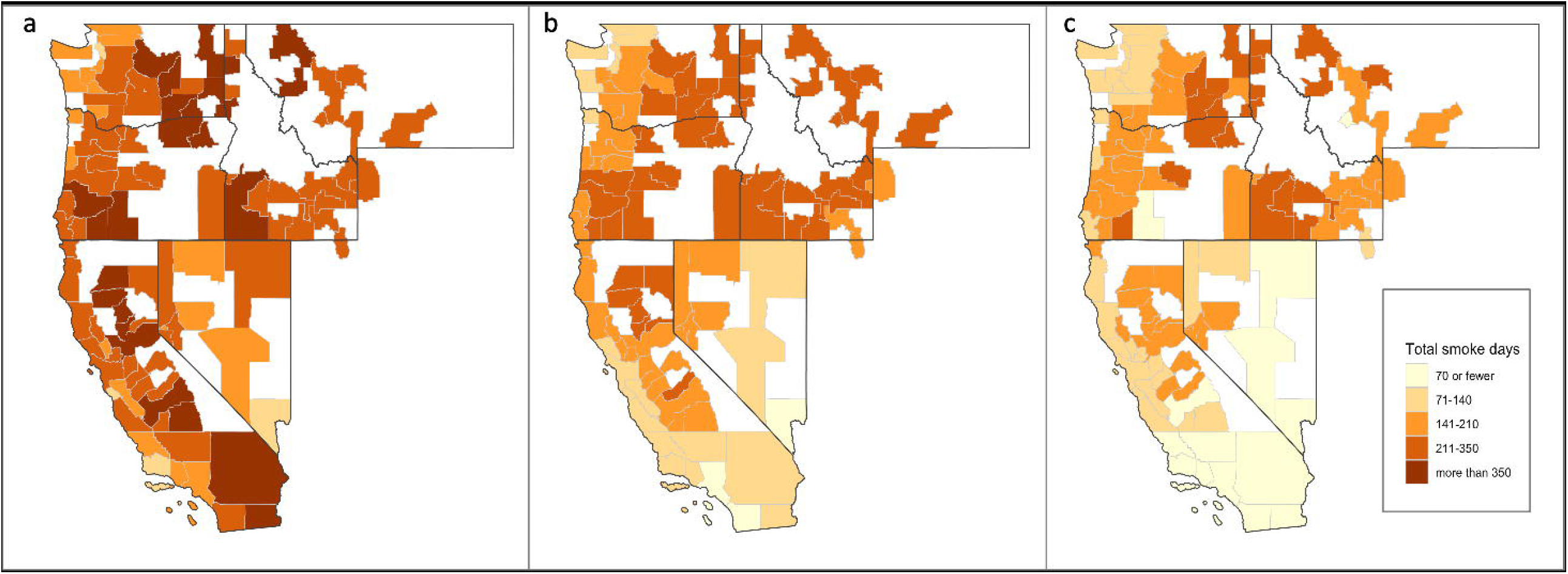
Spatial distribution of total number of smoke-days (2010-2016) at each MSA for the (a) 0%, (b) 25%, and (c) 50% thresholds. Thresholds are defined as the population-weighted percentage of zip codes within an MSA required to experience a smoke-day, in order to assign a smoke-day to the MSA.

Smoke-day exposures were linked to each liveborn child according to residential MSA at birth. For each child, the number of smoke-days were summed in each exposure period and divided by the number of weeks in the period to produce average weekly smoke-day estimates. This transformation allows comparison across periods with differing durations and across individuals with various GAs. Average weekly smoke-day estimates for each period were generated for each of the three exposure thresholds.

In the full cohort, we estimated the average weekly wildfire smoke-days for the following time periods: each trimester of gestation, and two consecutive 12-week periods after birth. First, second, and third trimesters (T1, T2, & T3) were assumed to be 280-197, 196-99, & 98-1 days before the estimated date of birth, respectively. In the full cohort, postnatal exposure periods, P1 and P2, were defined 1-84 days (1-12 weeks) and 85-168 days (13-24 weeks) after birth, respectively.

In the GA sub-cohort, we additionally used GA to more accurately estimate the average weekly smoke-days for the portion of each trimester completed (clarifying example in **Supplementary Material**). To distinguish between exposures obtained under the assumption of 40-week gestation (full cohort) and those obtained using estimated GA (GA sub-cohort), we use the suffix ‘-40w’ or ‘-GA’, respectively.

### Statistical Analyses

For each trimester and postnatal period, mixed effects Cox proportional hazards models were used to estimate the hazard ratio (HR) for the association between the weekly average smoke-days and each prescription outcome. Follow-up time was counted in weeks. The association between average weekly smoke exposure and first use of medications was estimated using the function below:

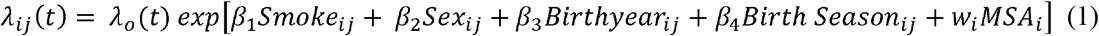

where *λ*_*ij*_(*t*) is the probability of the first filled respiratory prescription at time *t* for child *j* in the *i*^*th*^ MSA; the *MSA* into which a child was born is the random intercept, and *w*_*i*_ is vector of coefficients for the random intercept. *Smoke* is the average weekly number of smoke-days for the given trimester or postnatal period. In addition to the two models of postnatal smoke-day exposures, separate models were estimated for the six gestational smoke-day exposure periods (T1-40w, T2-40w, T3-40w, T1-GA, T2-GA, and T3-GA). A proxy for sex-specific hormones during development, *sex* was dichotomous (male/female).

*Birthyear* was included to adjust for time trends in both the propensity to prescribe medications and the increasing number of smoke-days. Socioeconomic status was not available in the MarketScan database within which all children were privately insured. The 4-category covariate *Birth Season*, defined as birth during Spring (March-May), Summer (June-August), Fall (September-November), or Winter (December-February), was included to adjust for potential confounding.

Results for each trimester exposure are presented both for full cohort (all eligible births, assuming a 40-week gestation) and for the GA sub-cohort (the subset of births with estimable gestational age).

Results for each postnatal exposure are presented for the full cohort only. Hazard ratios (HR (95% confidence interval(CI))) are estimated for a one day increase in weekly average smoke-days. Results are presented for all children as well as stratified on sex.

Though this analysis focuses on population-level effects rather than specific sub-populations or etiologies (work-in-progress), we recognize that preterm birth (PTB) that may drive our main findings. As such, we also stratified by PTB (defined as <37wks) within the GA-sub-cohort.

## RESULTS

Across the study region of 60 MSAs in six western states (U.S.A.), each MSA had between 31 (Walla Walla, WA) and 24,860 (Los Angeles-Long Beach-Glendale, California) births recorded in MarketScan during 2010-2016, with mean and median of 3,039 and 914 births per MSA, respectively (**Table S1**). Los Angeles, Portland, and Seattle contributed the MSAs with the greatest number of births. Of 182,387 eligible births with estimable birthdates (based on claims codes summarized in **Appendix 1**), 89,066 (48.8%) were female, and 113,154 (62.0%) also had an estimable GA (**Table 1**). The number of births per year ranged from 18,578 to 32,721, and the sex distribution remained similar across years. In the full cohort, Cetirizine hydrochloride (54.3%) and Mometasone furoate (34.8%) were the most frequently filled upper respiratory prescriptions; Albuterol sulfate (90.0%) was the most frequently filled lower respiratory prescription; Methylprednisolone, Prednisolone, or Prednisone (86.3%) was the most frequently filled systemic anti-inflammatory prescriptions (**Table S3**).

**Table 1.**
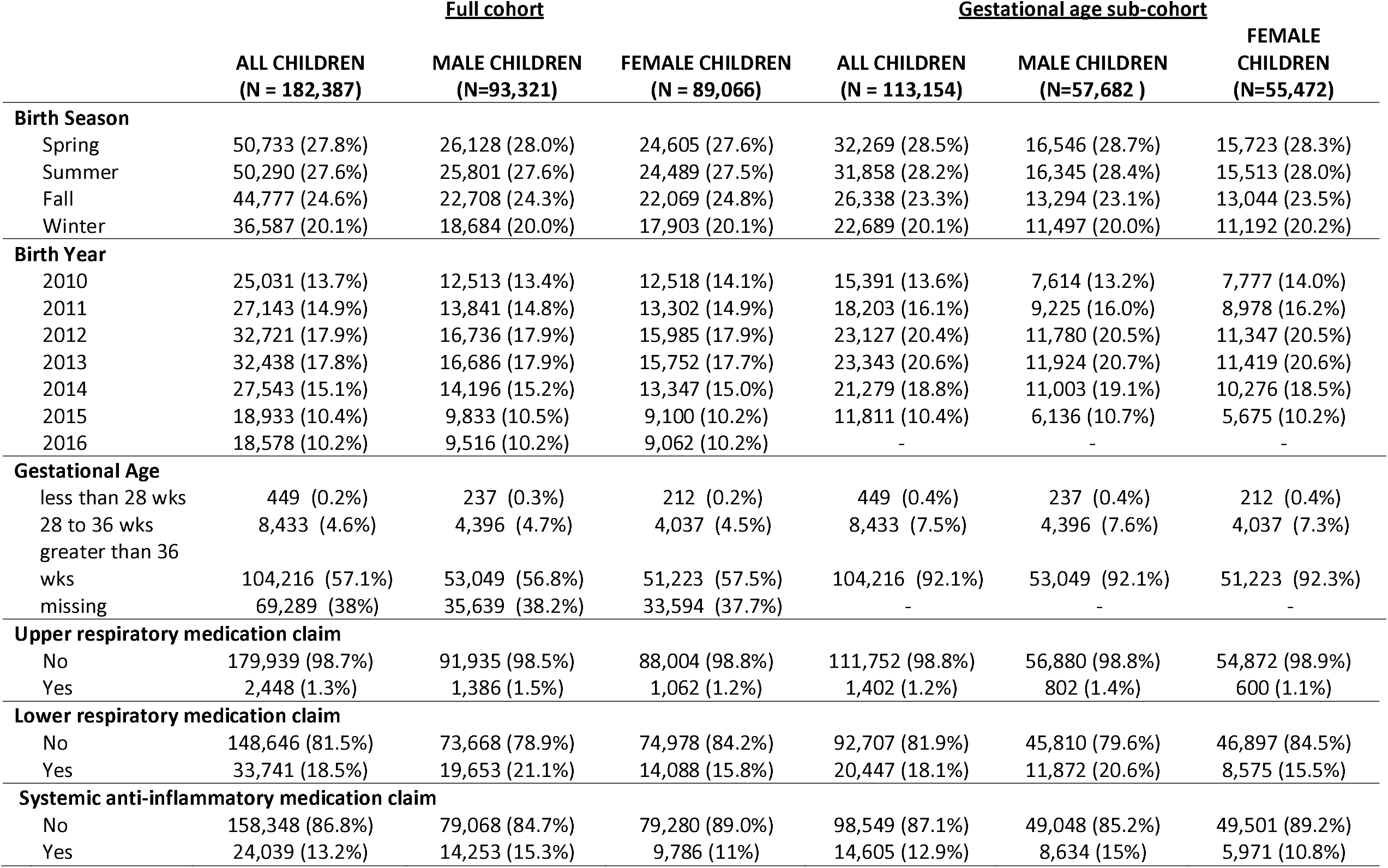
Cohort composition in the full cohort and the gestational age sub-cohort, overall and by sex.

|  | <u>Full cohort</u> |  |  | <u>Gestational age sub-cohort</u> |  |  |
| --- | --- | --- | --- | --- | --- | --- |
|  | **ALL CHILDREN**<br>**(N = 182,387)** | **MALE CHILDREN**<br>**(N=93,321)** | **FEMALE CHILDREN**<br>**(N = 89,066)** | **ALL CHILDREN**<br>**(N = 113,154)** | **MALE CHILDREN**<br>**(N=57,682 )** | **FEMALE**<br>**CHILDREN**<br>**(N=55,472)** |
| **Birth Season** |  |  |  |  |  |  |
| Spring | 50,733 (27.8%) | 26,128 (28.0%) | 24,605 (27.6%) | 32,269 (28.5%) | 16,546 (28.7%) | 15,723 (28.3%) |
| Summer | 50,290 (27.6%) | 25,801 (27.6%) | 24,489 (27.5%) | 31,858 (28.2%) | 16,345 (28.4%) | 15,513 (28.0%) |
| Fall | 44,777 (24.6%) | 22,708 (24.3%) | 22,069 (24.8%) | 26,338 (23.3%) | 13,294 (23.1%) | 13,044 (23.5%) |
| Winter | 36,587 (20.1%) | 18,684 (20.0%) | 17,903 (20.1%) | 22,689 (20.1%) | 11,497 (20.0%) | 11,192 (20.2%) |
| **Birth Year** |  |  |  |  |  |  |
| 2010 | 25,031 (13.7%) | 12,513 (13.4%) | 12,518 (14.1%) | 15,391 (13.6%) | 7,614 (13.2%) | 7,777 (14.0%) |
| 2011 | 27,143 (14.9%) | 13,841 (14.8%) | 13,302 (14.9%) | 18,203 (16.1%) | 9,225 (16.0%) | 8,978 (16.2%) |
| 2012 | 32,721 (17.9%) | 16,736 (17.9%) | 15,985 (17.9%) | 23,127 (20.4%) | 11,780 (20.5%) | 11,347 (20.5%) |
| 2013 | 32,438 (17.8%) | 16,686 (17.9%) | 15,752 (17.7%) | 23,343 (20.6%) | 11,924 (20.7%) | 11,419 (20.6%) |
| 2014 | 27,543 (15.1%) | 14,196 (15.2%) | 13,347 (15.0%) | 21,279 (18.8%) | 11,003 (19.1%) | 10,276 (18.5%) |
| 2015 | 18,933 (10.4%) | 9,833 (10.5%) | 9,100 (10.2%) | 11,811 (10.4%) | 6,136 (10.7%) | 5,675 (10.2%) |
| 2016 | 18,578 (10.2%) | 9,516 (10.2%) | 9,062 (10.2%) | - | - | - |
| **Gestational Age** |  |  |  |  |  |  |
| less than 28 wks | 449 (0.2%) | 237 (0.3%) | 212 (0.2%) | 449 (0.4%) | 237 (0.4%) | 212 (0.4%) |
| 28 to 36 wks | 8,433 (4.6%) | 4,396 (4.7%) | 4,037 (4.5%) | 8,433 (7.5%) | 4,396 (7.6%) | 4,037 (7.3%) |
| greater than 36 wks | 104,216 (57.1%) | 53,049 (56.8%) | 51,223 (57.5%) | 104,216 (92.1%) | 53,049 (92.1%) | 51,223 (92.3%) |
| missing | 69,289 (38%) | 35,639 (38.2%) | 33,594 (37.7%) | - | - | - |
| **Upper respiratory medication claim** |  |  |  |  |  |  |
| No | 179,939 (98.7%) | 91,935 (98.5%) | 88,004 (98.8%) | 111,752 (98.8%) | 56,880 (98.8%) | 54,872 (98.9%) |
| Yes | 2,448 (1.3%) | 1,386 (1.5%) | 1,062 (1.2%) | 1,402 (1.2%) | 802 (1.4%) | 600 (1.1%) |
| **Lower respiratory medication claim** |  |  |  |  |  |  |
| No | 148,646 (81.5%) | 73,668 (78.9%) | 74,978 (84.2%) | 92,707 (81.9%) | 45,810 (79.6%) | 46,897 (84.5%) |
| Yes | 33,741 (18.5%) | 19,653 (21.1%) | 14,088 (15.8%) | 20,447 (18.1%) | 11,872 (20.6%) | 8,575 (15.5%) |
| **Systemic anti-inflammatory medication claim** |  |  |  |  |  |  |
| No | 158,348 (86.8%) | 79,068 (84.7%) | 79,280 (89.0%) | 98,549 (87.1%) | 49,048 (85.2%) | 49,501 (89.2%) |
| Yes | 24,039 (13.2%) | 14,253 (15.3%) | 9,786 (11%) | 14,605 (12.9%) | 8,634 (15%) | 5,971 (10.8%) |

On average, children were exposed to less than one day of smoke per week in each exposure period; the mean trimester exposure ranged from 0.28 to 0.70 weekly smoke-days, and postnatal mean exposure ranged from 0.37 to 0.74, across the thresholds (**Table S4**). Descriptive statistics for the full cohort, the GA sub-cohort and those without GA were relatively comparable (**Table S5**).

### Postnatal period

In both the first (P1) and second (P2) postnatal periods, the models reflected an association between smoke-day exposure with earlier first use of upper respiratory medication (P1: HR=1.094 per 1-day increase in weekly smoke-day (1.005, 1.191); P2: HR=1.108 (1.016, 1.209)); **Figure 3, Table 2)**. In the P1 period, female children had the larger effect size and stronger association (HR=1.123 (0.994, 1.27)) as compared to males (HR=1.055 (0.940, 1.185)). In contrast, male children had a larger effect size (HR=1.128 (1.004, 1.266)) and stronger association in P2, as compared to female children (HR=1.072 (0.942, 1.22); **Figure 3, Table 2)**. Associations in primary models (i.e., using the 25% threshold) of lower respiratory as well as systemic anti-inflammatory outcomes were largely null (**Figure 3, Table 2**).

**Table 2.** Mixed effects cox model of smoke exposure during post-partum trimester at the 25% exposure threshold, in all eligible children and stratified by sex. P1 and P2 periods correspond to 0-12 and 13-24 weeks, respectively. Models are adjusted for birth-season, birthyear and, where applicable, sex, with a random intercept for MSA. All exposures are the average weekly number of smoke-days in the given post-partum period. RX = prescribed medication claim.

|  |  | ALL CHILDREN<br>(N = 182,387) | MALE CHILDREN<br>(N = 93,321) | FEMALE CHILDREN<br>(N = 89,066) |
| --- | --- | --- | --- | --- |
|  | Exposure | HR (95% CI) | HR (95% CI) | HR (95% CI) |
| Upper respiratory RX | P1 | 1.094 (1.005, 1.191) | 1.055 (0.940, 1.185) | 1.123 (0.994, 1.270) |
|  | P2 | 1.108 (1.016, 1.209) | 1.128 (1.004, 1.266) | 1.072 (0.942, 1.220) |
| Lower respiratory RX | P1 | 1.007 (0.983, 1.031) | 1.013 (0.982, 1.045) | 0.992 (0.955, 1.030) |
|  | P2 | 0.980 (0.955, 1.007) | 0.971 (0.938, 1.005) | 0.987 (0.947, 1.029) |
| Systemic Anti-inflammatory RX | P1 | 1.001 (0.975, 1.028) | 0.995 (0.961, 1.030) | 1.008 (0.966, 1.051) |
|  | P2 | 0.996 (0.969, 1.025) | 1.004 (0.968, 1.042) | 0.980 (0.938, 1.025) |

**Figure 3.**
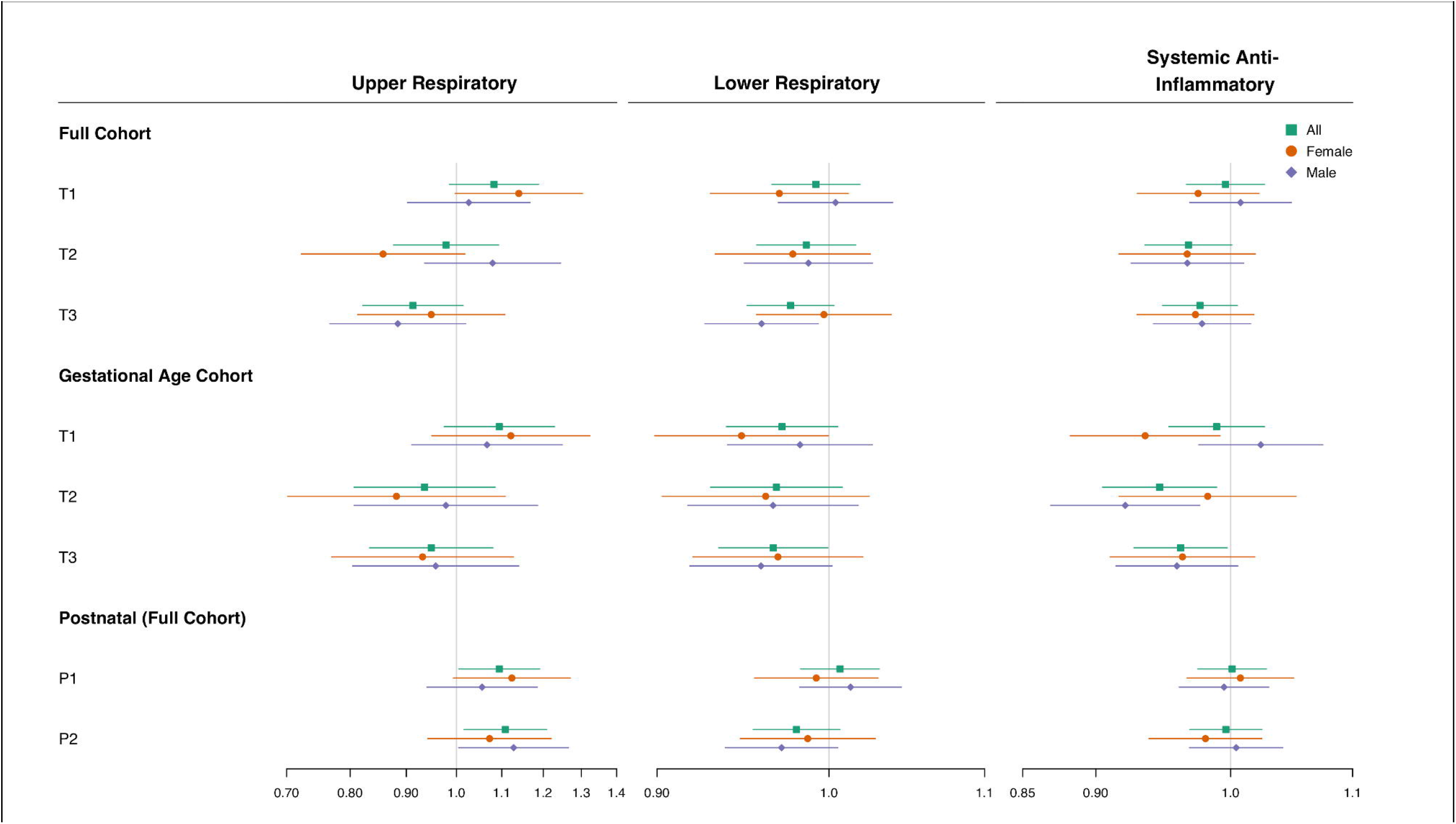
Hazard ratio and confidence intervals for the relationship of average weekly smoke-days and first use of prescription outcome, in each averaging period at the 25% exposure threshold, for all children (green square), and within strata of male children (purple diamonds) and female children (orange circle). The Cox proportional hazards models were adjusted for birth season, birth year, infant sex and included a random intercept for metropolitan statistical area; infant sex covariate was omitted in infant sex-stratified models. rx = prescribed medication claim; T1-T3 refer to the first-third trimesters; P1 and P2 refer to the first and second 12-week post-natal period.

### Gestational period

Positive associations with smoke exposure were observed only in the first trimester for the upper respiratory outcome. The 40w- and GA-exposure models produced HRs in the same direction with comparable magnitude, and with some variation in precision (**Figure 3, Table 3**).

**Table 3.**
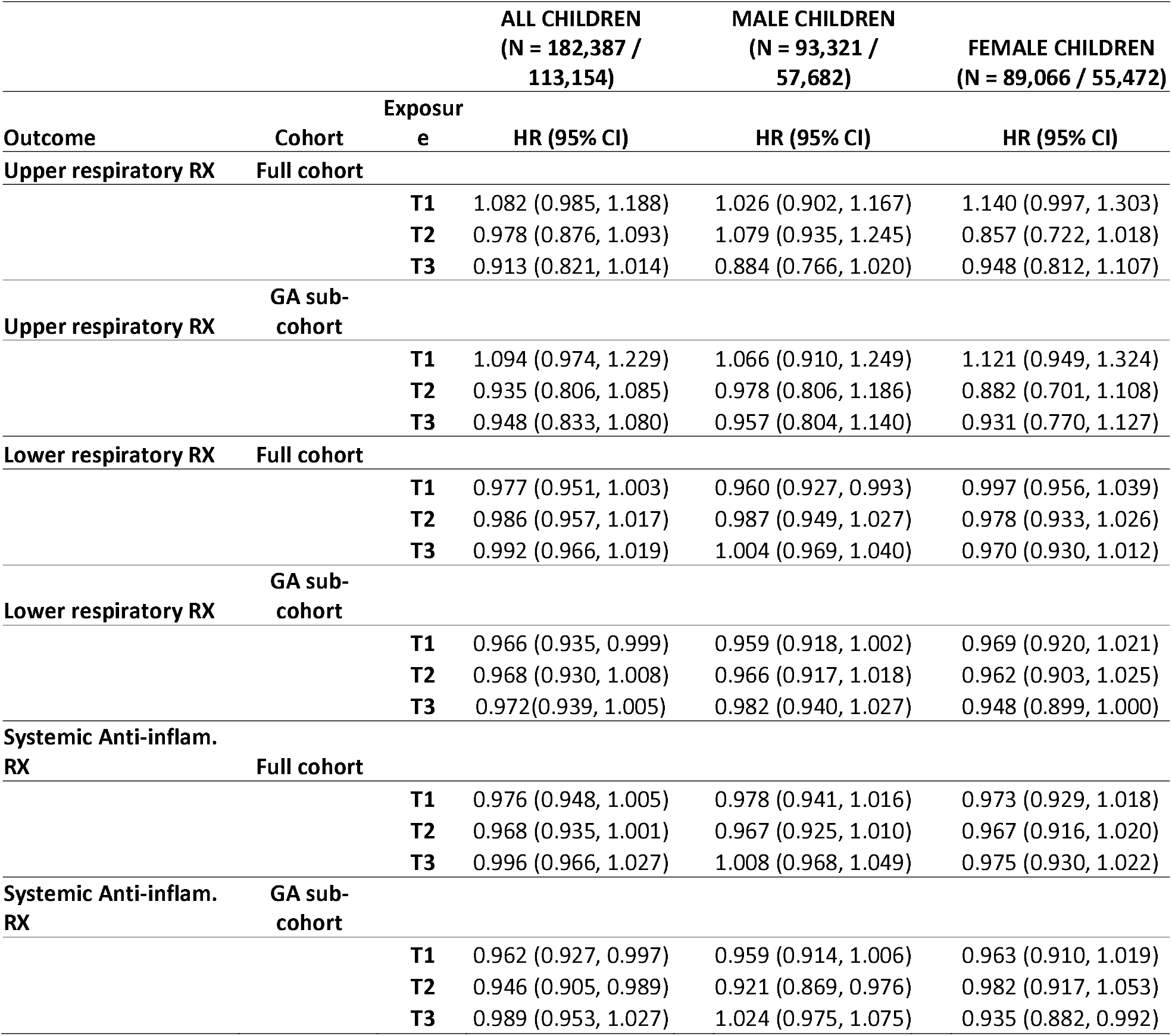
Mixed effects cox model results of smoke exposure during gestational periods at the 25% exposure threshold, in all eligible children and stratified by sex. Models are adjusted for birth season, birthyear and, where applicable, sex, with a random intercept for MSA. The first and second N values correspond to the entire cohort (full cohort) and the cohort portion whose gestational age estimate was available to estimate wildfire smoke-day exposure (GA sub-cohort), respectively. All exposures are the average weekly number of smoke-days in the given period. RX = prescribed medication claim.

#### Upper respiratory

In the primary model, the impact of smoke-days on the upper respiratory medications outcome varied by gestational period, with increased risk in the first trimester, and null or decreased risk in the second and third trimester (**Figure 3, Table 3**). In the first trimester, smoke-day exposure was associated with shorter times to first upper respiratory medication, among all (HR=1.082 (0.985, 1.188)), and female children (HR=1.140 (0.997, 1.303)) among the full cohort (T1-40w), but not in male children (HR=1.026 (0.902, 1.167); **Figure 3)**; these first trimester HRs are comparable in the GA sub-cohort (T1-GA), albeit with less precision in the 95%CI. For all other exposure windows, generally null associations were consistently observed in both the full cohort and the GA sub-cohort.

#### Lower respiratory

Smoke-days had a null or slightly inverse association across all three trimesters in the 40-week exposure modeled in both the full cohort and the GA sub-cohort (**Figure 3, Table 3**). In contrast to first trimester upper respiratory outcome findings, first trimester smoke-day exposure among the full cohort (T1-40w) was associated with delayed first lower respiratory medications among all (HR=0.977 (0.951, 1.003)) and male children (HR=0.960 (0.927, 0.993)), but not in female children (HR=0.997 (0.956, 1.039); **Figure 3**).

#### Systemic Anti-inflammatory

Second trimester smoke-day exposures in the GA sub-cohort (T2-GA) were inversely associated with longer time to the use of systemic anti-inflammatory medication in all (HR=0.946 (0.905, 0.989)) and male (HR=0.921 (0.869, 0.987)) children (**Figure 3, Table 3**). Similar, but smaller magnitude associations were observed in the first trimester (T1-GA) (All: HR=0.962 (0.927, 0.997); male: HR=0.959 (0.914, 1.006); **Figure 3)**). Interestingly, a similar protective association was found among female children in the third trimester (T3-GA) (HR=0.935 (0.892, 0.992)), but not among male children. In the full cohort, the exposures across all trimesters and groups produce consistent and similarly precise associations that are closer to the null than their counterparts in the GA sub-cohort.

### Sensitivity analyses

In the postnatal period, HRs and 95%CIs for both upper respiratory and systemic anti-inflammatory outcomes are approximately similar across the 0%, 25% and 50% exposure thresholds (**Figure S1**). For the lower respiratory outcome, however, results in the exposure sensitivity analyses are more variable across thresholds.

For all trimester outcomes, the various thresholds used to generate smoke-day exposure estimates generally had little effect on HR estimates, with some marginal changes in the confidence interval precision (**Figure S1**). HR did not markedly differ between term births and PTBs.

## DISCUSSION

Using private insurance claims data, we conducted a time-to-event analysis to estimate the association between developmental exposure to wildfire smoke and first use of respiratory medication. We found an association between smoke exposure during the first trimester, and both postnatal periods with shorter time to first use of upper respiratory medication; these results were robust across exposure definitions. This positive association between wildfire exposure and upper respiratory medication used was larger for female children in the first trimester and the first 12-week postnatal period, while it was stronger for male children in the 13-24 postnatal week period. These stratified results may suggest sex-specific windows of vulnerability.

Additionally, for both systemic anti-inflammatory medication and lower respiratory medication, protective effects of wildfire exposure were observed during most gestational windows. Most prominently, we observed a shorter time to first lower respiratory medication use among male children and all children during the first trimester; this finding was relatively consistent across cohort specification (i.e., full cohort vs GA sub-cohort). Similarly, all children and male children had shorter times to first systemic anti-inflammatory medication with increased smoke exposure during the first and second trimesters; this was true also of girls in the first trimester. Whether the mechanism for these observed protective effects is an artifact of observational epidemiology or a causal biological finding cannot be ascertained in this dataset.

Our findings suggest a complex epidemiological relationship between gestational exposure to wildfire smoke and time to first symptoms requiring respiratory prescription use. The juxtaposition of below-null HRs during gestation with evidence in the literature suggesting that ambient air pollution is not protective indicates that this analysis may suffer from selection bias (as do most perinatal epidemiological studies (34)). In administrative datasets, these analyses are necessarily limited to live births, and do not capture pregnancy loss. If the *in utero* response to wildfire smoke exposure is strong, restricting the cohort to live births could lead to attrition of susceptible pregnancies and to an *in utero*-outcome association which is biased to appear less harmful or even protective. Indeed, non-human primate work shows pregnancy loss is associated with wildfire smoke exposure (35).

The differing exposure pathways in the gestational and postnatal periods could also account for the discrepancy in findings between these two developmental windows. Broadly, possible pathways for air pollutants to exert influence on a fetus include altered placentation or placenta-mediated processes, and direct fetal exposure. Recent work showed that PM can deposit in placental tissue and, to a lesser extent, pass through the placenta to fetal capillaries (36–38). The increased tidal volume in the birthing parent as well as the increased respiratory rate in young children may result in a larger dose of air pollutants (39,40). Should this increased deposition of wildfire smoke pollutants in the birthing parent impair placentation, there may be increased fetal loss as observed in non-human primates and mentioned above (35). In the surviving fetuses, however, our inconsistent or null findings in the gestational period as compared to positive upper respiratory postnatal findings, could indicate that direct exposure to nasal epithelia, as occurs in the postnatal period, produces a more prominent and detectable increase in use of respiratory medications.

Previous studies of PM exposures suggest specific respiratory conditions that could result from wildfire smoke exposure, and may or may not require prescriptions treatment. For example, developmental exposures to PM or biomass burning are associated with early life deficits in lung function (41–43), and increased susceptibility to acute respiratory infection (ARI) or infant bronchiolitis (44,45). While ARI is often treated with prescription medication in young children, subclinical deficits in lung function associated with developmental air pollution exposure may not always be identified and treated with prescription medication unless they lead to respiratory infection (21,43,46–51). It is possible that study cohort members experienced respiratory symptoms that were transient, sub-clinical or well-managed with non-prescription medication, and thus were not captured in this analysis blunting any potential association.

Because lung development occurs in phases starting in early gestation and continuing into adolescence, damage in early life may have lasting negative effects. As PM is a component of wildfire smoke exposure, the mechanisms by which such negative effects occur likely include a combination of oxidative stress and inflammation, which may further induce physiological (52–54) or epigenetic changes in the birthing parent and/or offspring (55,56). Early gestational exposure may lead to abnormal cellular differentiation or branching due to a disruption of essential redox signaling (52), while later exposure may induce structural or functional abnormalities (53,54). In non-human primates, early life exposure to woodsmoke has been linked to reduced lung function metrics (24) which may be due to epigenetic changes to respiratory tissues (57).

To our knowledge, this is the first analysis describing the impact of exposure to wildfire smoke during development on prescription-treated respiratory conditions in human children. There are several additional strengths in this analysis. We were able to represent multiple exposure scenarios by using various thresholds for assigning smoke-days to an MSA. For example, because the definition of smoke-day is based on the appearance of a plume from satellite data, adjacent areas may indeed be exposed to a lower level of smoke that, while not visible, is still impactful; this scenario is captured by the 0% threshold. We also explored refining exposure estimate using GA. Despite losing 38% of the dataset by including claims-based estimates of GA, exaggerated effect estimates observed in the GA sub-cohort suggest that accounting for GA captures important aspects of exposure relative to fetal development that the assumption of 40-week gestation does not. The use of prescription claims data more accurately captures medication usage than diagnosis or prescription delivery captured in electronic medical records. Further the thorough categorization of these medications by two physicians ensures that they are meaningfully classified. Finally, we were the first to leverage two widely used datasets to answer our research question: well-vetted wildfire exposure data and private claims data able to capture prescription usage.

First use of respiratory prescription medication as described in a private insurance claims database serves as an indicator of respiratory health and vulnerability; this approach is not without limitations. The MarketScan Commercial Claims and Encounters database is limited by omission of those who are not on private insurance and those living in rural areas; this could make our results less generalizable to other groups because of socioeconomic factors. While the Marketscan claims database allows the examination of a large population distributed over a large wildfire-impacted region, the availability of individual-level confounders, such as race, SES, and maternal or second-hand smoking, is unfortunately limited. As observed with other air pollutant exposures (58), SES could modify the effect of smoke exposure on respiratory health. Relatedly, this analysis assumes that all participants are exposed to the same proportion of outdoor air pollution (e.g., that they spend the same amount of time outside and all have the same/no indoor air purification systems), which may contribute to the potential for exposure bias differential on SES or other related factors.

Imprecision in the estimates of residence may lead to exposure misclassification, though this impact is expected to be small given the size of a MSA in which exposures are defined as uniform due to the spatial coarseness of the data. MSA at birth provides a relatively coarse estimate of exposure which could be improved upon by using a dataset which includes more granular residential information. We also assumed that the MSA at time of birth was the same during the entirety of gestation; while likely not universally true, the impact of this assumption is expected to be small with potential bias toward the null (59,60). Additionally, the coarseness of gestational age available in the ICD-9 coding system, wherein codes are available at 2-week intervals between 24-37 weeks (inclusive), somewhat limits the accuracy of our exposure estimates in the GA sub-cohort; such exposure misclassification may also induce bias in the effect estimates. Finally, some respiratory outcomes may be lost by claims data, as it is only able to provide information on conditions treated by prescription for those who have access to necessary care.

The modest differences in associations by sex found in this analysis contribute to growing evidence supporting sex-differential effects of wildfire smoke exposure. There are well-documented sex differences in lung development and asthma (61), including stress-induced oxidative response among fetuses (62). Noted sex-specific reductions in lung function in non-human primates following wildfire smoke exposure during infancy (24) may be exacerbated by prenatal PM exposure (21).

Overall, these findings suggest that first trimester and postnatal wildfire smoke exposure is associated with shorter time to the first upper respiratory medication usage, while gestational wildfire smoke exposure across trimesters is associated with longer time to systemic anti-inflammatory respiratory prescription usage. This research supports the growing body of literature indicating that wildfire smoke exposure in early life poses a health risk for pediatric populations, and adds granularity to the current understanding of sex-specific and trimester-specific effects of in utero wildfire smoke exposure.

Clinicians may consider discussing the potential benefits of reducing exposure using evidenced-based measures such as observing air quality index alerts, placing air purifiers in their home, and wearing face masks to reduce particulate exposures. Future work should investigate specific clinical outcomes, such as acute respiratory infection, and utilize finer spatial and temporal granularity to refine estimates of wildfire smoke exposure. This work also highlights the continued need to evaluate measures to protect against wildfire smoke during key developmental stages.

## Data Availability

All wildfire smoke satellite data are publicly available through NASA at https://www.ospo.noaa.gov/Products/land/hms.html
Office of Housing and Urban Development data is available at https://www.huduser.gov/portal/datasets/usps_crosswalk.html#data
Claims data are proprietary and may be obtained form IBM MarketScan (https://www.ibm.com/products/marketscan-research-databases).

https://www.huduser.gov/portal/datasets/usps_crosswalk.html#data

https://www.ibm.com/products/marketscan-research-databases

## Declarations

### Authors contributions

RD, MER, and IJ conceived of the study question. AGR, CWC, and IJ obtained study funding. RD, CK, YX, and BSS built the dataset and carried out statistical analyses. KR, AGR, and CW-C advised on statistical analyses and cohort development from claims data. RD classified medications as to the outcome and AC and MH reviewed these classifications. RD, CK, and HVJ drafted the manuscript.

### Consent for publication

All authors revised the manuscript critically for intellectual content, gave final approval of the version to be published, and agreed to be accountable for all aspects of the work.

### Competing interests

The authors declare they have no competing interests to disclose.

### Funding

Supported by the Environmental Protection Agency (EPA) Cooperative Agreement (U.S. EPA CR83578501); HVJ was supported in part by an appointment to the ORISE participant research program supported by an interagency agreement between EPA and DOE. CK was supported in part by a training grant from the National Institute of Environmental Health Sciences [T32ES007018]. BSS was supported by the Genetic Epidemiology of Heart, Lung, and Blood Traits Training Grant [T32HL129982].

### Disclaimer

The views expressed in this article are those of the authors and do not necessarily represent the views or policies of the U.S. Environmental Protection Agency.

### Availability of data and materials

IBM MarketScan® Research Databases, a proprietary claims dataset, on which these analyses are based are available through IBM (https://www.ibm.com/products/marketscan-research-databases). The environmental data from the NOAA Hazards Mapping System that support the conclusions of this article are available in the U.S. EPA Environmental Dataset Gateway (https://www.ospo.noaa.gov/Products/land/hms.html - maps).

### Ethics approval

This study (IRB No:20-2817) was evaluated and approved by the Institutional Review Board at the University of North Carolina at Chapel Hill.

## Acknowledgements

The authors would like to acknowledge Michele Jonnson-Funk, Virginia Pate, and Tom Luben for their support and guidance in this developing this work.

## Abbreviations

WSE: Wildfire smoke exposure
MSA: Metropolitan Statistical Area
PM: Particulate matter
PM_2.5_: fine fraction of particulate matter
MarketScan: IBM MarketScan® Commercial Claims and Encounters Research Database
ICD: International Classification of Disease
ZCTA: ZIP Code Tabulation Area
MSTFMDS: mode of administration
THERCLS: therapeutic drug classes
THERGRP: therapeutic drug groups
GA: Gestational Age
T1, T2, T3: 1^st^, 2^nd^ and 3^rd^ trimesters, respectively
P1 and P2: 1^st^ and 2^nd^ consecutive 12-week postnatal period, respectively
HR: hazard ratio
95% CI: 95% confidence interval
PTB: pre-term birth

## Notes

### Competing Interest Statement

The authors have declared no competing interest.

### Funding Statement

This study was funded by the Environmental Protection Agency (EPA) Cooperative Agreement (U.S. EPA CR83578501). HVJ was supported in part by an appointment to the ORISE participant research program supported by an interagency agreement between EPA and DOE. CK was supported in part by a training grant from the National Institute of Environmental Health Sciences [T32ES007018]. BSS was supported by the Genetic Epidemiology of Heart, Lung, and Blood Traits Training Grant [T32HL129982].

### Author Declarations

Institutional Review Board at the University of North Carolina at Chapel Hill gave ethical approval for this work.

### Summary of Updates

Revisions include clarifications and minor corrections that reflect feedback from reviewers in previous journal submissions. This preprint may also be deposited at Environmental Health Journal, at the request of journal editors.

## References

1. National Interagency Fire Center. Total Wildland Fires and Acres (1983-2021) [Internet]. [cited 2022 Mar 24]. (Statistics). Available from: https://www.nifc.gov/fire-information/statistics/wildfires

2. Fann N, Alman B, Broome RA, Morgan GG, Johnston FH, Pouliot G, et al. The health impacts and economic value of wildland fire episodes in the U.S.: 2008–2012. Sci Total Environ. 2018 Jan 1;610–611:802–9.

3. Liu JC, Peng RD. The impact of wildfire smoke on compositions of fine particulate matter by ecoregion in the Western US. J Expo Sci Environ Epidemiol. 2019 Nov;29(6):765–76.

4. Lelieveld J, Pozzer A, Pöschl U, Fnais M, Haines A, Münzel T. Loss of life expectancy from air pollution compared to other risk factors: a worldwide perspective. Cardiovasc Res. 2020 Sep 1;116(11):1910–7.

5. Künzli N, Avol E, Wu J, Gauderman WJ, Rappaport E, Millstein J, et al. Health effects of the 2003 Southern California wildfires on children. Am J Respir Crit Care Med. 2006 Dec 1;174(11):1221–8.

6. Reid CE, Maestas MM. Wildfire smoke exposure under climate change: impact on respiratory health of affected communities. Curr Opin Pulm Med. 2019 Mar;25(2):179–87.

7. Rebuli ME, Speen AM, Martin EM, Addo KA, Pawlak EA, Glista-Baker E, et al. Wood Smoke Exposure Alters Human Inflammatory Responses to Viral Infection in a Sex-Specific Manner. A Randomized, Placebo-controlled Study. Am J Respir Crit Care Med. 2019 Apr 15;199(8):996–1007.

8. Sigsgaard T, Forsberg B, Annesi-Maesano I, Blomberg A, Bølling A, Boman C, et al. Health impacts of anthropogenic biomass burning in the developed world. Eur Respir J. 2015 Dec;46(6):1577–88.

9. DeFlorio-Barker S, Crooks J, Reyes J, Rappold AG. Cardiopulmonary Effects of Fine Particulate Matter Exposure among Older Adults, during Wildfire and Non-Wildfire Periods, in the United States 2008-2010. Environ Health Perspect. 2019 Mar;127(3):37006.

10. Wettstein ZS, Hoshiko S, Fahimi J, Harrison RJ, Cascio WE, Rappold AG. Cardiovascular and Cerebrovascular Emergency Department Visits Associated With Wildfire Smoke Exposure in California in 2015. J Am Heart Assoc. 2018;7(8):e007492.

11. Rappold AG, Stone SL, Cascio WE, Neas LM, Kilaru VJ, Carraway MS, et al. Peat bog wildfire smoke exposure in rural North Carolina is associated with cardiopulmonary emergency department visits assessed through syndromic surveillance. Environ Health Perspect. 2011 Oct;119(10):1415–20.

12. Haikerwal A, Akram M, Del Monaco A, Smith K, Sim MR, Meyer M, et al. Impact of Fine Particulate Matter (PM2.5) Exposure During Wildfires on Cardiovascular Health Outcomes. J Am Heart Assoc. 2015 Jul 15;4(7):e001653.

13. Morgan G, Sheppeard V, Khalaj B, Ayyar A, Lincoln D, Jalaludin B, et al. Effects of bushfire smoke on daily mortality and hospital admissions in Sydney, Australia. Epidemiol Camb Mass. 2010 Jan;21(1):47–55.

14. Delfino RJ, Brummel S, Wu J, Stern H, Ostro B, Lipsett M, et al. The relationship of respiratory and cardiovascular hospital admissions to the southern California wildfires of 2003. Occup Environ Med. 2009 Mar;66(3):189–97.

15. Mishra V. Indoor air pollution from biomass combustion and acute respiratory illness in preschool age children in Zimbabwe. Int J Epidemiol. 2003 Oct 1;32(5):847–53.

16. Oloyede IP, Ekrikpo UE, Ekanem EE. Lung Function Indices of Children Exposed to Wood Smoke in a Fishing Port in South-South Nigeria. J Trop Pediatr. 2013 Oct 1;59(5):399–402.

17. Fullerton DG, Suseno A, Semple S, Kalambo F, Malamba R, White S, et al. Wood smoke exposure, poverty and impaired lung function in Malawian adults. Int J Tuberc Lung Dis. 2011 Mar 1;15(3):391–8.

18. García-Sancho MC, García-García L, Báez-Saldaña R, Ponce-de-León A, Sifuentes-Osornio J, Bobadilla-del-Valle M, et al. Indoor pollution as an occupational risk factor for tuberculosis among women: a population-based, gender oriented, case-control study in Southern Mexico. :8.

19. Pérez-Padilla R, Pérez-Guzmán C, Báez-Saldaña R, Torres-Cruz A. Cooking with biomass stoves and tuberculosis: a case control study. Int J Tuberc Lung Dis Off J Int Union Tuberc Lung Dis. 2001 May;5(5):441–7.

20. Wright RJ, Brunst KJ. Programming of respiratory health in childhood: influence of outdoor air pollution. Curr Opin Pediatr. 2013 Apr;25(2):232–9.

21. Hsu HL, Chiu YM, Coull BA, Schwartz J, Lee A, Wright RO, et al. Prenatal Particulate Air Pollution and Asthma Onset in Urban Children. Identifying Sensitive Windows and Sex Differences. Am J Respir Crit Care Med. 2015 Nov;192(9):1052–9.

22. Stapleton A, Casas M, García J, García R, Sunyer J, Guerra S, et al. Associations between pre- and postnatal exposure to air pollution and lung health in children and assessment of CC16 as a potential mediator. Environ Res. 2022 Mar 1;204:111900.

23. Schultz ES, Hallberg J, Bellander T, Bergström A, Bottai M, Chiesa F, et al. Early-Life Exposure to Traffic-related Air Pollution and Lung Function in Adolescence. Am J Respir Crit Care Med. 2016 Jan 15;193(2):171–7.

24. Black C, Gerriets JE, Fontaine JH, Harper RW, Kenyon NJ, Tablin F, et al. Early Life Wildfire Smoke Exposure Is Associated with Immune Dysregulation and Lung Function Decrements in Adolescence. Am J Respir Cell Mol Biol. 2017 May 1;56(5):657–66.

25. Li L, Jick S, Breitenstein S, Hernandez G, Michel A, Vizcaya D. Pulmonary arterial hypertension in the USA: an epidemiological study in a large insured pediatric population. Pulm Circ. 2017 Mar;7(1):126–36.

26. Li Q, Andrade SE, Cooper WO, Davis RL, Dublin S, Hammad TA, et al. Validation of an algorithm to estimate gestational age in electronic health plan databases. Pharmacoepidemiol Drug Saf. 2013 May;22(5):524–32.

27. Butler AM, Layton JB, Li D, Hudgens MG, Boggess KA, McGrath LJ, et al. Predictors of Low Uptake of Prenatal Tetanus Toxoid, Reduced Diphtheria Toxoid, and Acellular Pertussis Immunization in Privately Insured Women in the United States. Obstet Gynecol. 2017 Apr;129(4):629–37.

28. Truven Health Analytics/IBM Watson Health. IBM Micromedex RedBook [Internet]. 2021. Available from: https://www.micromedexsolutions.com

29. Up To Date. Up To Date. 2021.

30. Rolph GD, Draxler RR, Stein AF, Taylor A, Ruminski MG, Kondragunta S, et al. Description and Verification of the NOAA Smoke Forecasting System: The 2007 Fire Season. Weather Forecast. 2009 Apr 1;24(2):361–78.

31. Schroeder W, Ruminski M, Csiszar I, Giglio L, Prins E, Schmidt C, et al. Validation analyses of an operational fire monitoring product: The Hazard Mapping System. Int J Remote Sens. 2008 Oct 20;29(20):6059–66.

32. OFFICE OF POLICY DEVELOPMENT AND RESEARCH (PD&R). HUD USPS ZIP CODE CROSSWALK FILES [Internet]. U.S. Department of Housing and Urban Development; [cited 2021 Nov 12]. Available from: https://www.huduser.gov/portal/datasets/usps_crosswalk.html#data

33. Wilson, Ron, Din, Alexander. Understanding and Enhancing the U.S. Department of Housing and Urban Development’s ZIP Code Crosswalk Files. Cityscape J Policy Dev Res. 2018;20(2):277–94.

34. Snowden JM, Bovbjerg ML, Dissanayake M, Basso O. The curse of the perinatal epidemiologist: inferring causation amidst selection. Curr Epidemiol Rep. 2018 Dec;5(4):379–87.

35. Willson BE, Gee NA, Willits NH, Li L, Zhang Q, Pinkerton KE, et al. Effects of the 2018 Camp Fire on birth outcomes in non-human primates: Case-control study. Reprod Toxicol. 2021 Oct 1;105:128–35.

36. Bongaerts E, Nawrot TS, Van Pee T, Ameloot M, Bové H. Translocation of (ultra)fine particles and nanoparticles across the placenta; a systematic review on the evidence of in vitro, ex vivo, and in vivo studies. Part Fibre Toxicol. 2020 Nov 2;17(1):56.

37. Bongaerts E, Aengenheister L, Dugershaw BB, Manser P, Roeffaers MBJ, Ameloot M, et al. Label-free detection of uptake, accumulation, and translocation of diesel exhaust particles in ex vivo perfused human placenta. J Nanobiotechnology. 2021 May 17;19(1):144.

38. Engelen L. Translocation of airborne black carbon particles from mother to unborn child: an ENVIRONAGE birth cohort study. :29.

39. Chang J, Streitman D. Physiologic Adaptations to Pregnancy. Neurol Clin. 2012 Aug 1;30(3):781–9.

40. Fleming S, Thompson M, Stevens R, Heneghan C, Plüddemann A, Maconochie I, et al. Normal ranges of heart rate and respiratory rate in children from birth to 18 years: a systematic review of observational studies. Lancet. 2011 Mar 19;377(9770):1011–8.

41. Bai H, Capitanio JP, Miller LA, Clougherty JE. Social status and susceptibility to wildfire smoke among outdoor-housed female rhesus monkeys: A natural experiment. Heliyon. 2021 Nov 1;7(11):e08333.

42. Schultz ES, Gruzieva O, Bellander T, Bottai M, Hallberg J, Kull I, et al. Traffic-related Air Pollution and Lung Function in Children at 8 Years of Age: A Birth Cohort Study. Am J Respir Crit Care Med. 2012 Dec 15;186(12):1286–91.

43. Jedrychowski WA, Perera FP, Spengler JD, Mroz E, Stigter L, Flak E, et al. Intrauterine exposure to fine particulate matter as a risk factor for increased susceptibility to acute broncho-pulmonary infections in early childhood. Int J Hyg Environ Health. 2013 Jul;216(4):395–401.

44. Odo DB, Yang IA, Knibbs LD. A Systematic Review and Appraisal of Epidemiological Studies on Household Fuel Use and Its Health Effects Using Demographic and Health Surveys. Int J Environ Res Public Health. 2021 Feb 3;18(4):1411.

45. Karr C, Demers P, Koehoorn M. Influence of ambient air pollutant sources on clinical encounters for infant bronchiolitis. Am J Respir Crit Care Med. 2009;180:995–1001.

46. Seeni I, Ha S, Nobles C, Liu D, Sherman S, Mendola P. Air pollution exposure during pregnancy: maternal asthma and neonatal respiratory outcomes. Ann Epidemiol. 2018 Sep 1;28(9):612-618.e4.

47. Rosa MJ, Just AC, Kloog I, Pantic I, Schnaas L, Lee A, et al. Prenatal particulate matter exposure and wheeze in Mexican children: Effect modification by prenatal psychosocial stress. Ann Allergy Asthma Immunol. 2017 Sep 1;119(3):232-237.e1.

48. Chiu YHM, Coull BA, Sternthal MJ, Kloog I, Schwartz J, Cohen S, et al. Effects of prenatal community violence and ambient air pollution on childhood wheeze in an urban population. J Allergy Clin Immunol. 2014 Mar 1;133(3):713-722.e4.

49. Soh SE, Goh A, Teoh OH, Godfrey KM, Gluckman PD, Shek LPC, et al. Pregnancy Trimester-Specific Exposure to Ambient Air Pollution and Child Respiratory Health Outcomes in the First 2 Years of Life: Effect Modification by Maternal Pre-Pregnancy BMI. Int J Environ Res Public Health. 2018 May;15(5):996.

50. Latzin P, Röösli M, Huss A, Kuehni CE, Frey U. Air pollution during pregnancy and lung function in newborns: a birth cohort study. Eur Respir J. 2009 Mar 1;33(3):594–603.

51. Jedrychowski WA, Perera FP, Maugeri U, Mroz E, Klimaszewska-Rembiasz M, Flak E, et al. Effect of prenatal exposure to fine particulate matter on ventilatory lung function of preschool children of non-smoking mothers. Paediatr Perinat Epidemiol. 2010;24(5):492–501.

52. Land SC, Wilson SM. Redox Regulation of Lung Development and Perinatal Lung Epithelial Function. Antioxid Redox Signal. 2005 Jan;7(1–2):92–107.

53. Kajekar R. Environmental factors and developmental outcomes in the lung. Pharmacol Ther. 2007 May 1;114(2):129–45.

54. Korten I, Ramsey K, Latzin P. Air pollution during pregnancy and lung development in the child. Paediatr Respir Rev. 2017 Jan 1;21:38–46.

55. Dugershaw BB, Aengenheister L, Hansen SSK, Hougaard KS, Buerki-Thurnherr T. Recent insights on indirect mechanisms in developmental toxicity of nanomaterials. Part Fibre Toxicol. 2020 Jul 11;17(1):31.

56. Veras MM, de Oliveira Alves N, Fajersztajn L, Saldiva P. Before the first breath: prenatal exposures to air pollution and lung development. Cell Tissue Res. 2017 Mar 1;367(3):445–55.

57. Brown AP, Cai L, Laufer BI, Miller LA, LaSalle JM, Ji H. Long-term effects of wildfire smoke exposure during early life on the nasal epigenome in rhesus macaques. Environ Int. 2022 Jan 1;158:106993.

58. O’ Lenick CR, Chang HH, Kramer MR, Winquist A, Mulholland JA, Friberg MD, et al. Ozone and childhood respiratory disease in three US cities: evaluation of effect measure modification by neighborhood socioeconomic status using a Bayesian hierarchical approach. Environ Health. 2017 Apr 5;16(1):36.

59. Chen L, Bell EM, Caton AR, Druschel CM, Lin S. Residential mobility during pregnancy and the potential for ambient air pollution exposure misclassification. Environ Res. 2010 Feb 1;110(2):162–8.

60. Pennington AF, Strickland MJ, Klein M, Zhai X, Russell AG, Hansen C, et al. Measurement error in mobile source air pollution exposure estimates due to residential mobility during pregnancy. J Expo Sci Environ Epidemiol. 2017 Sep;27(5):513–20.

61. Carey MA, Card JW, Voltz JW, Arbes SJ, Germolec DR, Korach KS, et al. It’s all about sex: gender, lung development and lung disease. Trends Endocrinol Metab. 2007 Oct 1;18(8):308–13.

62. Minghetti L, Greco A, Zanardo V, Suppiej A. Early-life sex-dependent vulnerability to oxidative stress: the natural twining model. J Matern Fetal Neonatal Med. 2013 Feb;26(3):259–62.

