## Supplemental Figures, Tables, and Appendices for "Wildfire smoke exposure and early childhood respiratory health: a study of prescription claims data"

Supplemental Materials: Wildfire smoke exposure and early childhood respiratory outcomes: an examination of private prescription claims data

Radhika Dhingra ^1*^, Corinna Keeler ^2^, Brooke S. Staley ^2^, Hanna V. Jardel ^2,3^, Cavin Ward-Caviness ^3^, Meghan E. Rebuli ^4,5^, Yuzhi Xi ^1^, Kristen Rappazzo ^3^, Michelle Hernandez ^4^, Anne Chelminski ^3^, Ilona Jaspers ^4,5^, Ana G. Rappold ^3^

^1^ Department of Environmental Sciences and Engineering, Gillings School of Global Public Health, University of North Carolina at Chapel Hill, Chapel Hill, NC, USA.

^2^ Department of Epidemiology, Gillings School of Global Public Health, University of North Carolina at Chapel Hill, Chapel Hill, NC, USA.

^3^ United States Environmental Protection Agency, Center for Public Health and Environmental Assessment, Research Triangle Park, NC, USA.

^4^ Department of Pediatrics, School of Medicine, University of North Carolina at Chapel Hill, Chapel Hill, NC, USA.

^5^ Center for Environmental Medicine, Asthma, and Lung Biology, University of North Carolina at Chapel Hill, Chapel Hill, NC, USA.

***Corresponding Author:** Radhika Dhingra, PhD, Department of Environmental Science and Engineering, Gillings School of Global Public Health, University of North Carolina, 135 Dauer Drive, C.B 7431, Chapel Hill, NC 27599. E-mail address:

### Supplemental Figures

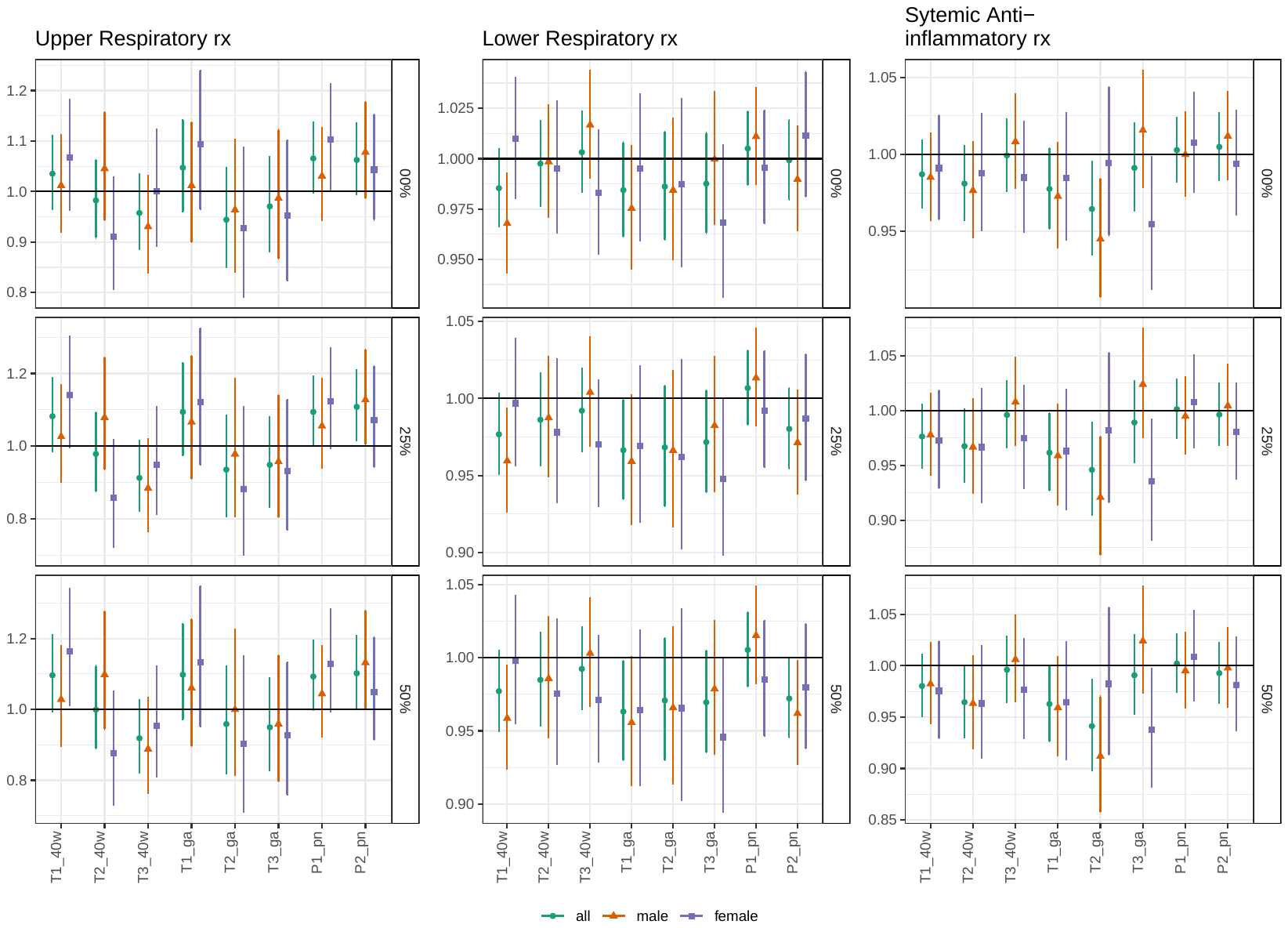

**Figure S1.** Hazard ratios and 95% confidence intervals Cox proportional hazards models across exposure thresholds. rx = prescribed medication claim.

### Supplemental Tables

**Table S1**. Number of yearly births in study region Metropolitan statistical areas (MSA).

| **MSA Code** | **Name** | **2010** | **2011** | **2012** | **2013** | **2014** | **2015** | **2016** | **TOTAL** |
| --- | --- | --- | --- | --- | --- | --- | --- | --- | --- |
| 10540 | Albany, OR Metro Area | -- | -- | -- | -- | 102 | 114 | 77 | 293 |
| 12540 | Bakersfield-Delano, CA Metro Area (2010-2012); Bakersfield, CA Metro Area (2013-2016) | 342 | 334 | 332 | 319 | 190 | 88 | 96 | 1701 |
| 13380 | Bellingham, WA Metro Area | 47 | 54 | 44 | 55 | 60 | 68 | 43 | 371 |
| 13460 | Bend, OR Metro Area (2010-2012);  Bend-Redmond, OR Metro Area (2013-2016) | 118 | 134 | 132 | 117 | 129 | 127 | 99 | 856 |
| 13740 | Billings, MT Metro Area | 303 | 291 | 344 | 305 | 92 | 64 | 58 | 1457 |
| 14260 | Boise City-Nampa, ID Metro Area (2010-2012);  Boise City, ID Metro Area (2013-2016) | 404 | 437 | 1363 | 1272 | 1399 | 1316 | 1336 | 7527 |
| 14740 | Bremerton-Silverdale, WA Metro Area | 36 | 41 | 72 | 65 | 73 | 49 | 47 | 383 |
| 16180 | Carson City, NV Metro Area | 43 | 21 | 21 | 20 | 18 | 9 | 11 | 143 |
| 17020 | Chico, CA Metro Area | 138 | 119 | 156 | 145 | 79 | 34 | 31 | 702 |
| 17660 | Coeur d'Alene, ID Metro Area | 65 | 46 | 260 | 233 | 262 | 270 | 261 | 1397 |
| 18700 | Corvallis, OR Metro Area | 39 | 31 | 37 | 37 | 32 | 32 | 25 | 233 |
| 20940 | El Centro, CA Metro Area | 107 | 77 | 63 | 38 | 23 | 11 | 12 | 331 |
| 21660 | Eugene-Springfield, OR Metro Area (2010-2012); Eugene,OR Metro Area (2013-2016) | 156 | 151 | 165 | 157 | 155 | 124 | 105 | 1013 |
| 23420 | Fresno, CA Metro Area | 386 | 384 | 416 | 419 | 312 | 183 | 161 | 2261 |
| 24420 | Grants Pass, OR Metro Area | -- | -- | -- | -- | 19 | 19 | 15 | 53 |
| 24500 | Great Falls, MT Metro Area | 173 | 170 | 187 | 144 | 14 | 21 | 8 | 717 |
| 25260 | Hanford-Corcoran, CA Metro Area | 29 | 35 | 55 | 42 | 12 | 14 | 11 | 198 |
| 26820 | Idaho Falls, ID Metro Area | 61 | 110 | 408 | 345 | 405 | 380 | 333 | 2042 |
| 28420 | Kennewick-Richland, WA Metro Area (2010-2012); Kennewick-Richland, WA Metro Area (2013-2016) | 70 | 81 | 155 | 124 | 145 | 126 | 115 | 816 |
| 29820 | Las Vegas-Paradise, NV Metro Area (2010-2012);  Las Vegas-Henderson-Paradise, NV Metro Area (2013-2016) | 1508 | 1393 | 1251 | 1411 | 1264 | 1030 | 1088 | 8945 |
| 30300 | Lewiston, ID-WA Metro Area | 11 | 14 | 53 | 53 | 56 | 60 | 54 | 301 |
| 30860 | Logan, UT-ID Metro Area | 56 | 82 | 158 | 140 | 147 | 146 | 136 | 865 |
| 31020 | Longview, WA Metro Area | 31 | 21 | 45 | 31 | 45 | 33 | 29 | 235 |
| 31084 | Los Angeles-Long Beach-Glendale, CA Metropolitan Division | 3571 | 3896 | 4651 | 4538 | 4079 | 2066 | 2059 | 24860 |
| 31460 | Madera-Chowchilla, CA Metro Area (2010-2012); Madera, CA Metro Area (2013-2016) | 47 | 62 | 46 | 53 | 48 | 28 | 36 | 320 |
| 32780 | Medford, OR Metro Area | 102 | 115 | 98 | 75 | 59 | 61 | 40 | 550 |
| 32900 | Merced, CA Metro Area | 99 | 123 | 119 | 113 | 89 | 50 | 44 | 637 |
| 33540 | Missoula, MT Metro Area | 217 | 219 | 201 | 198 | 24 | 37 | 22 | 918 |
| 33700 | Modesto, CA Metro Area | 197 | 222 | 208 | 250 | 168 | 131 | 130 | 1306 |
| 34580 | Mount Vernon-Anacortes, WA Metro Area | 28 | 38 | 47 | 47 | 43 | 36 | 44 | 283 |
| 34900 | Napa, CA Metro Area | 63 | 58 | 66 | 59 | 35 | 12 | 12 | 305 |
| 36084 | Oakland-Hayward-Berkeley, CA Metropolitan Division | 1363 | 1341 | 1653 | 1520 | 1631 | 949 | 1000 | 9457 |
| 36500 | Olympia, WA Metro Area (2010-2012);  Olympia-Tumwater, WA Metro Area (2013-2013) | 99 | 119 | 149 | 104 | 101 | 87 | 64 | 723 |
| 37100 | Oxnard-Thousand Oaks-Ventura, CA Metro Area | 495 | 571 | 613 | 596 | 469 | 264 | 253 | 3261 |
| 38540 | Pocatello, ID Metro Area | 70 | 47 | 225 | 221 | 220 | 187 | 166 | 1136 |
| 38900 | Portland-Vancouver-Hillsboro, OR-WA Metro Area | 1881 | 2303 | 2546 | 2498 | 2407 | 2397 | 1706 | 15738 |
| 39820 | Redding, CA Metro Area | 98 | 85 | 87 | 63 | 55 | 43 | 31 | 462 |
| 39900 | Reno-Sparks, NV Metro Area (2010-2012);  Reno, NV Metro Area (2013-2016) | 307 | 235 | 262 | 267 | 210 | 153 | 165 | 1599 |
| 40140 | Riverside-San Bernardino-Ontario, CA Metro Area | 1291 | 1474 | 1627 | 1490 | 1215 | 688 | 711 | 8496 |
| 40900 | Sacramento--Arden-Arcade--Roseville, CA Metro Area (2010-2012); Sacramento--Roseville--Arden-Arcade, CA Metro Area (2013-2016) | 1464 | 1671 | 1698 | 1587 | 1041 | 897 | 829 | 9187 |
| 41420 | Salem, OR Metro Area | 286 | 331 | 345 | 385 | 356 | 372 | 240 | 2315 |
| 41500 | Salinas, CA Metro Area | 295 | 331 | 294 | 313 | 183 | 94 | 84 | 1594 |
| 41740 | San Diego-Carlsbad-San Marcos, CA Metro Area (2010-2012); San Diego-Carlsbad, CA Metro Area (2013-2016) | 1136 | 1162 | 1466 | 1649 | 1399 | 709 | 1269 | 8790 |
| 41884 | San Francisco-Redwood City-South San Francisco, CA Metropolitan Division | 1166 | 1207 | 1517 | 1707 | 1792 | 1112 | 1137 | 9638 |
| 41940 | San Jose-Sunnyvale-Santa Clara, CA Metro Area | 1417 | 1546 | 1859 | 2229 | 2141 | 878 | 925 | 10995 |
| 42020 | San Luis Obispo-Paso Robles, CA Metro Area (2010-2012); San Luis Obispo-Paso Robles-Arroyo Grande, CA Metro Area (2013-2016) | 255 | 273 | 270 | 270 | 117 | 42 | 33 | 1260 |
| 42044 | Santa Ana-Anaheim-Irvine, CA Metropolitan Division | 1485 | 1755 | 2010 | 1949 | -- | -- | -- | 7199 |
| 42060 | Santa Barbara-Santa Maria-Goleta, CA Metro Area | 147 | 154 | 203 | 224 | -- | -- | -- | 728 |
| 42100 | Santa Cruz-Watsonville, CA Metro Area | 191 | 215 | 249 | 239 | 198 | 90 | 92 | 1274 |
| 42220 | Santa Rosa-Petaluma, CA Metro Area (2010-2012); Santa Rosa, CA Metro Area (2013-2016) | 149 | 144 | 135 | 146 | 146 | 80 | 90 | 890 |
| 42644 | Seattle-Bellevue-Everett, WA Metropolitan Division | 1741 | 1978 | 2621 | 2671 | 2870 | 2183 | 2214 | 16278 |
| 44060 | Spokane, WA Metro Area (2010-2012); Spokane-Spokane Valley, WA Metro Area (2013-2016) | 257 | 273 | 322 | 199 | 194 | 187 | 202 | 1634 |
| 44700 | Stockton, CA Metro Area (2010-2012); Stockton-Lodi, CA Metro Area (2013-2016) | 265 | 302 | 354 | 342 | 259 | 135 | 134 | 1791 |
| 45104 | Tacoma-Lakewood, WA Metropolitan Division | 324 | 378 | 524 | 488 | 538 | 346 | 398 | 2996 |
| 46700 | Vallejo-Fairfield, CA Metro Area | 146 | 166 | 174 | 138 | 127 | 81 | 77 | 909 |
| 47300 | Visalia-Porterville, CA Metro Area | 104 | 146 | 171 | 125 | 90 | 56 | 54 | 746 |
| 47460 | Walla Walla, WA Metro Area | -- | -- | -- | -- | 11 | 7 | 13 | 31 |
| 48300 | Wenatchee-East Wenatchee, WA Metro Area (2010-2012); Wenatchee, WA Metro Area (2013-2016) | 19 | 26 | 44 | 51 | 50 | 43 | 47 | 280 |
| 49420 | Yakima, WA Metro Area | 42 | 45 | 70 | 71 | 77 | 58 | 57 | 420 |
| 49700 | Yuba City, CA Metro Area | 91 | 106 | 80 | 91 | 68 | 56 | 49 | 541 |

**Table S2**. Overview of generic and brand names for Medications associated with each outcome.

| **Drug class** | **Medication** | **Brand name examples** |
| --- | --- | --- |
| **Upper Respiratory Medications** | | |
| **Inhaled nasal steroids** | Fluticasone (nasal) | Flonase, Flonase Sensimist |
|  | Budesonide (nasal) | Rhinocort |
|  | Mometasone (nasal) | Nasonex |
|  | Beclomethasone (nasal) | Beconase |
|  | Fluticasone fuorate (nasal) | Veramyst |
| **Singulair** | Montelukast | Singulair |
| **Antihistamine** | Cetirizine | Zyrtec |
|  | Fexofenadine | Allegra |
|  | Loratidine | Claritin |
|  | Levocetirizine | Xyzal |
| **Lower Respiratory Medications** | | |
| **Short-acting Beta Agonist (SABA)** | Albuterol (salbutamol) | AccuNeb, Proair HFA, ProAir RespiClick, Proventil HFA, Ventolin HFA, Vospire ER, Metaprotenol |
|  | Levalbuterol | Xopenex HFA |
|  | Pirbuterol | Maxair |
| **Injectable asthma biologics** | Omalizumab | Xolair |
| **Long-acting Bronchodilators combinations (LABA)  + inhaled corticosteroids (ICS))** | Fluticasone propionate+ salmeterol | Advair |
|  | mometasone furoate + formoterol fumarate dihydrate | Dulera |
|  | Fluticasone fuorate + vilanterol | Breo Ellipta |
|  | budesonide & formoterol | Symbicort |
| **Inhaled steroids** | Fluticasone | Flovent |
|  | Budesonide | Pulmicort |
|  | Mometasone | Asthmanex |
|  | Beclomethasone | Qvar |
|  | Fluticasone fuorate | Arnuity Ellipta |
| **Singulair** | Montelukast | singulair |
| **Systemic Anti-inflammatory Medications** | | |
| **Oral or injectable Steroids** | Prednisolone |  |
|  | Methylprednisolone |  |
|  | Prednisone |  |
|  | Dexamethasone |  |

**Table S3.** Frequency of prescription medication by drug class. Only generic names are presented here. Frequency in each drug class may sum to more than 100%, as more than one medication in a drug class may be prescribed in a single claims record.

|  | | **N** | **%** |
| --- | --- | --- | --- |
| **Upper Respiratory prescription medication** | |  |  |
|  | Azelastine hydrochloride | 20 | 0.8% |
|  | Beclomethasone dipropionate | 11 | 0.4% |
|  | Budesonide | 7 | 0.3% |
|  | Cetirizine hydrochloride | 1,329 | 54.3% |
|  | Fexofenadine hydrochloride | 4 | 0.2% |
|  | Fluticasone furoate | 93 | 3.8% |
|  | Levocetirizine dihydrochloride | 83 | 3.4% |
|  | Loratadine | 70 | 2.9% |
|  | Mometasone furoate | 852 | 34.8% |
| **Lower Respiratory prescription medication** | |  |  |
|  | Albuterol Sulfate | 30,360 | 90.0% |
|  | Albuterol Sulfate/Ipratropium Bromide | 43 | 0.1% |
|  | Beclomethasone Dipropionate | 417 | 1.2% |
|  | Budesonide | 2,128 | 6.3% |
|  | Budesonide/Formoterol | 2 | 0.01% |
|  | Levalbuterol hydrochloride | 1,815 | 5.4% |
|  | Levalbuterol tartrate | 362 | 1.1% |
|  | Mometasone furoate | 6 | 0.02% |
|  | Montelukast sodium | 29 | 0.1% |
| **Systemic Anti-inflammatory prescription medication** | | | |
|  | Dexamethasone | 3,507 | 14.6% |
|  | Methylprednisolone, Prednisolone, or Prednisone | 20,741 | 86.3% |

**Table S4.** Weekly average smoke-day exposure for each averaging period. Unit is average weekly number of smoke-days during given period. RX = prescribed medication claim.

|  | **Full cohort  (N = 182,387)** | | **Gestational age sub-cohort  (N = 113,154)** | | | |
| --- | --- | --- | --- | --- | --- | --- |
|  | **Mean (SD)** | **Median** | | **Mean (SD)** | | **Median** |
| **T1, Threshold = 0%** | 0.70 (0.90) | 0.33 | | 0.70 (0.90) | | 0.33 |
| **T2, Threshold = 0%** | 0.60 (0.79) | 0.29 | | 0.57 (0.76) | | 0.29 |
| **T3, Threshold = 0%** | 0.67 (0.87) | 0.29 | | 0.66 (0.88) | | 0.31 |
| **T1, Threshold = 25%** | 0.39 (0.63) | 0.08 | | 0.37 (0.61) | | 0.08 |
| **T2, Threshold = 25%** | 0.32 (0.53) | 0.07 | | 0.29 (0.50) | | 0.07 |
| **T3, Threshold = 25%** | 0.38 (0.62) | 0.07 | | 0.37 (0.63) | | 0.08 |
| **T1, Threshold = 50%** | 0.34 (0.59) | 0.08 | | 0.32 (0.57) | | 0.08 |
| **T2, Threshold = 50%** | 0.28 (0.50) | 0.07 | | 0.26 (0.47) | | 0.07 |
| **T3, Threshold = 50%** | 0.34 (0.59) | 0.07 | | 0.33 (0.60) | | 0.08 |
| **P1, Threshold = 0%** | 0.74 (0.93) | 0.33 | | - | - | |
| **P2, Threshold = 0%** | 0.72 (0.95) | 0.33 | | - | - | |
| **P1, Threshold = 25%** | 0.42 (0.67) | 0.17 | | - | - | |
| **P2, Threshold = 25%** | 0.41 (0.68) | 0.08 | | - | - | |
| **P1, Threshold = 50%** | 0.37 (0.64) | 0.08 | | - | - | |
| **P2, Threshold = 50%** | 0.37 (0.64) | 0.08 | | - | - | |

**Table S5.** Descriptive statistics in the full cohort and those with (i.e., GA sub-cohort) and without gestational age estimates.

|  |  | **Full cohort N = 182,387** | **Cohort with gestational age  N = 113,154** | **Cohort without gestational age N = 69,233** |
| --- | --- | --- | --- | --- |
| **Birth Season** | | |  |  |
|  | Spring | 50,733 (27.8%) | 32,269 (28.5%) | 18,464 (26.7%) |
|  | Summer | 50,290 (27.6%) | 31,858 (28.2%) | 18,432 (26.6%) |
|  | Fall | 44,777 (24.6%) | 26,338 (23.3%) | 18,439 (26.6%) |
|  | Winter | 36,587 (20.1%) | 22,689 (20.1%) | 13,898 (20.1%) |
| **Birth Year*** | | |  |  |
|  | 2010 | 25,031 (13.7%) | 15,391 (13.6%) | 9,640 (13.9%) |
|  | 2011 | 27,143 (14.9%) | 18,203 (16.1%) | 8,940 (12.9%) |
|  | 2012 | 32,721 (17.9%) | 23,127 (20.4%) | 9,594 (13.9%) |
|  | 2013 | 32,438 (17.8%) | 23,343 (20.6%) | 9,095 (13.1%) |
|  | 2014 | 27,543 (15.1%) | 21,279 (18.8%) | 6,264 (9.0%) |
|  | 2015 | 18,933 (10.4%) | 11,811 (10.4%) | 7,122 (10.3%) |
|  | 2016 | 18,578 (10.2%) | - | 18,578 (26.8%) |
| **Upper respiratory medication** | | |  |  |
|  | No | 179,939 (98.7%) | 111,752 (98.8%) | 68,187 (98.5%) |
|  | Yes | 2,448 (1.3%) | 1,402 (1.2%) | 1,046 (1.5%) |
| **Lower respiratory medication** | | |  |  |
|  | No | 148,646 (81.5%) | 92,707 (81.9%) | 55,939 (80.8%) |
|  | Yes | 33,741 (18.5%) | 20,447 (18.1%) | 13,294 (19.2%) |
| **Systemic anti-inflammatory medication** | | |  |  |
|  | No | 158,348 (86.8%) | 98,549 (87.1%) | 59,799 (86.4%) |
|  | Yes | 24,039 (13.2%) | 14,605 (12.9%) | 9,434 (13.6%) |
| * The transition of ICD-9 to ICD-10 codes occurring in late 2015. | | | | |

### Appendix 1. Algorithm for estimation of birthdate.

Adapted from (Li et al. 2017; [DOI: 10.1086/690007](https://pubmed.ncbi.nlm.nih.gov/28680572/)).

Estimation of birthdates for live born infants born between 2010 and 2016 was based on a search for ICD-9 and ICD-10 birth codes (**Table below**).

1. We identified all records with MarketScan recorded birth year between 2010-2016.
2. We examined the date of the first birth code. If the date of the first birth code matched the year of birth recorded in MarketScan, we assumed the date of the first recorded birth code was the birthdate.

Note, we chose not include the less stringent criteria presented in the Li algorithm, as the week-level precision was essential to exposure estimation.

| **ICD-9 Code** | **ICD-9 Description** | **ICD-10 Code** | **ICD-10 Description** |
| --- | --- | --- | --- |
| V3000 | Single Liveborn | Z3800 | Single liveborn infant, delivered vaginally |
| V3001 | Single Liveborn, cesarean | Z3801 | Single liveborn infant, delivered by cesarean |
| V301 | Single Liveborn, born outside hospital, hospitalized | Z381 | Single liveborn infant, born outside hospital |
| V302 | Single Liveborn, born outside hospital, not hospitalized | Z381 | Single liveborn infant, born outside hospital |
| V3100 | Twin Liveborn, Mate Liveborn | Z3830 | Twin liveborn infant, delivered vaginally |
| V3101 | Twin Liveborn, Mate Liveborn, delivered by cesarean | Z3831 | Twin liveborn infant, delivered by cesarean |
| V311 | Twin Liveborn, Mate Liveborn, born outside hospital, hospitalized | Z384 | Twin liveborn infant, born outside hospital |
| V312 | Twin Liveborn, Mate Liveborn, born outside hospital, not hospitalized | Z384 | Twin liveborn infant, born outside hospital |
| V3200 | Twin Liveborn, Mate Stillborn | Z3830 | Twin liveborn infant, delivered vaginally |
| V3201 | Twin Liveborn, Mate Stillborn, delivered by cesarean | Z3831 | Twin liveborn infant, delivered by cesarean |
| V321 | Twin Liveborn, Mate Stillborn, born outside hospital, hospitalized | Z384 | Twin liveborn infant, born outside hospital |
| V322 | Twin Liveborn, Mate Stillborn, born outside hospital, not hospitalized | Z384 | Twin liveborn infant, born outside hospital |
| V3300 | Twin Liveborn | Z3830 | Twin liveborn infant, delivered vaginally |
| V3301 | Twin Liveborn, delivered by cesarean | Z3831 | Twin liveborn infant, delivered by cesarean |
| V331 | Twin Liveborn, born outside hospital, hospitalized | Z384 | Twin liveborn infant, born outside hospital |
| V332 | Twin Liveborn, born outside hospital, not hospitalized | Z384 | Twin liveborn infant, born outside hospital |
| V3400 | Other Multiple liveborn, Mates Liveborn | Z3861 | Triplet liveborn infant, delivered vaginally |
| V3401 | Other Multiple liveborn, Mates Liveborn, delivered by caesarean | Z3869 | Other multiple liveborn infant, delivered by cesarean |
| V341 | Other Multiple liveborn, Mates Liveborn, born outside hospital, hospitalized | Z387 | Other multiple liveborn infant, born outside hospital |
| V342 | Other Multiple liveborn, Mates Liveborn, born outside hospital, not hospitalized | Z387 | Other multiple liveborn infant, born outside hospital |
| V3500 | Other Multiple liveborn, Mates Stillborn | Z3868 | Other multiple liveborn infant, delivered vaginally |
| V3501 | Other Multiple liveborn, Mates Stillborn, delivered by caesarean | Z3869 | Other multiple liveborn infant, delivered by cesarean |
| V351 | Other Multiple liveborn, Mates Stillborn, born outside hospital, hospitalized | Z387 | Other multiple liveborn infant, born outside hospital |
| V352 | Other Multiple liveborn, Mates Stillborn, born outside hospital, not hospitalized | Z387 | Other multiple liveborn infant, born outside hospital |
| V3600 | Other Multiple liveborn, Mates Liveborn and Stillborn | Z3868 | Other multiple liveborn infant, delivered vaginally |
| V3601 | Other Multiple liveborn, Mates Liveborn and Stillborn, delivered by caesarean | Z3869 | Other multiple liveborn infant, delivered by cesarean |
| V361 | Other Multiple liveborn, Mates Liveborn and Stillborn, born outside hospital, hospitalized | Z387 | Other multiple liveborn infant, born outside hospital |
| V362 | Other Multiple liveborn, Mates Liveborn and Stillborn, born outside hospital, not hospitalized | Z387 | Other multiple liveborn infant, born outside hospital |
| V3700 | Other Multiple liveborn | Z3868 | Other multiple live-born infant, delivered vaginally |
| V3701 | Other Multiple liveborn, delivered by caesarean | Z3869 | Other multiple liveborn infant, delivered by cesarean |
| V371 | Other Multiple liveborn, born outside hospital, hospitalized | Z387 | Other multiple liveborn infant, born outside hospital |
| V372 | Other Multiple liveborn, born outside hospital, not hospitalized | Z387 | Other multiple liveborn infant, born outside hospital |

### Appendix 2. Algorithm for Estimation of Gestational Age.

Adapted from (Butler et al. 2017; [DOI: 10.1097/AOG.0000000000001927](https://pubmed.ncbi.nlm.nih.gov/28277354/)).

To estimate gestational age at birth, we linked infant to the mother (birthing parent) and used a hierarchical algorithm using diagnosis and procedure codes from the infant and/or the mother (birthing parent).

1. We examined the infant diagnosis codes for evidence of premature delivery, that indicate either gestational age or gestational weight.
   a. If the infant had a diagnosis code for premature delivery with a reported range of gestational ages (ICD-9-CM codes 765.21-765.29), we imputed the upper value of the range category (e.g., an ICD-9-CM code of 76523, described as 25-26 completed weeks of gestation, is imputed as gestational age = 26 weeks).
   b. If, in the absence of reported gestational age, the infant had a diagnosis code for prematurity or extreme prematurity with birth weights reported (ICD-9-CM codes 765.01-765.09, 765.11-765.19), we imputed gestational age based on standard gestational growth charts.
   c. If, in the absence of reported weights or ages, the infant had a generic code for prematurity, we imputed gestational age as 35 weeks, or 28 weeks for a general code for extreme prematurity
2. If no information was obtained in steps 1a-c from the infant, and the mother (birthing parent) had a code for prematurity (ICD-9-CM code 644.2 or 644.21), we imputed gestational age as 35 weeks.
3. If the mother (birthing parent) had a code for long gestation (ICD-9-CM code 645.1, 645.11, 645.2, 645.21), we imputed gestational age as 42 weeks.
4. If there were no codes for prematurity or long gestation from the infant or mother, we imputed standard values depending on the birth outcome:
   a. For liveborn singletons, we imputed gestational age as 39 weeks.
   b. For multiple births, we imputed gestational age as 35 weeks.
5. In the case where both pre- and post-maturity codes were observed in the records of a mother (birthing parent) – infant dyad, the gestational age for those few records were recorded as not available.

### References

Butler AM, Layton JB, Li D, Hudgens MG, Boggess KA, McGrath LJ, et al. 2017. Predictors of Low Uptake of Prenatal Tetanus Toxoid, Reduced Diphtheria Toxoid, and Acellular Pertussis Immunization in Privately Insured Women in the United States. Obstet Gynecol 129:629–637; doi:10.1097/AOG.0000000000001927.

Li L, Jick S, Breitenstein S, Hernandez G, Michel A, Vizcaya D. 2017. Pulmonary arterial hypertension in the USA: an epidemiological study in a large insured pediatric population. Pulm Circ 7:126–136; doi:10.1086/690007.
